# Evaluation of p16/Ki- 67 cytology as triage test for high-risk HPV-positive women in a “see and treat strategy”

**DOI:** 10.1101/2025.01.26.25321149

**Authors:** Calleb George Onyango, Lilian Ogonda, Bernard Guyah

**Affiliations:** Department of Biomedical Science and Technology, Maseno University, Kisumu, Kenya

**Keywords:** cervical cancer screening, hr-HPV, p16/Ki67 cytology, VIA

## Abstract

**Background:** Although high risk human papilloma virus DNA (hr-HPV-DNA) test is the primary tool for cervical cancer screening, with visual inspection with acetic acid (VIA) serving as a triage test where Pap cytology is not available, the low intra-observer agreement associated with VIA means its reliability is limited and a more efficient test is still required. The aim of this study was to compare the performance of p16/Ki-67 cytology with VIA in the detection of cervical precancer and the feasibility as an alternative triage in the “*see and treat strategy”*.

**Methods:** In a hospital-based cross-sectional study, we utilized stored provider-collected specimens from a previous study of women referred with cervical abnormalities to a tertiary hospital in Kisumu County, Kenya from February 2021 to November 2023. Specimens were tested with both Xpert and p16/ki-67 Immunostain. All hr-HPV positive women with cervical lesions were triaged using VIA and p16/Ki- 67 cytology. CIN2 or worse (≥CIN2) were defined as the clinical end points.

**Results:** The p16/ Ki- 67 Immunostaining showed a statistically significant higher sensitivity (84.6% vs. 59.0%%), specificity (44.0% vs. 62.0%), positive predictive value (28.2% vs. 28.8%) and negative predictive value (91.7% vs. 85.3%) compared to VIA examination.

**Conclusion:** The p16/Ki-67 immunostaining for the detection of ≥CIN2 has shown high sensitivity and high negative predictive value in our study, which is comparable to several previous findings; implying that the assay is superior to VIA in identifying ≥CIN2 and can serve as an alternative tool for triaging primary HPV-positive women in the current “see and treat” strategy.

## Introduction

Cervical cancer (CC) incidence and mortality continue to rise despite every effort made to contain the disease leading to major global health imbalance that require urgent attention by all stakeholders [1]. Globally, CC ranks 8th in incidence amongst women with an estimated 660,000 new cases and 348,000 deaths annually [2]. The disease burden is more pronounced especially in sub-Saharan Africa (SSA) owing to high prevalence of HIV in the region [3]; more so in Eastern Africa where age-standardized incidence rate is above 40.0 per 100,000 women [4].

Uterine cervical intraepithelial neoplasia (CIN) is caused primarily by persistent infection with Human Papilloma Virus (HPV), especially of types 16 and 18 [2]. The virus penetrates the basal layers of epithelial cells through micro-abrasion of the transformation zone of the cervix using two oncogenes E6 and E7 [5] but which also interact with and inhibit various cell cycle-regulating protein such as retinoblastoma gene product pRB, and p53 protein; resulting in cervical dyskaryosis following a series of proteolytic degradation. Increasing expression of these viral oncogenes in dysplastic cervical cells is reflected by increased co-expression of p16INK4a and Ki-67 [6]. In Kenya, HPV-DNA test has been recommended as the primary screening tool for cervical cancer (CC) with subsequent triage using VIA where Pap cytology is not available owing to the fact that majority HPV infections are transient and do not cause the disease [7]. However, the low intra-observer agreement associated with VIA suggest that the reliability of VIA is so limited that its integration in treatment plan should be employed with a lot of caution [8]; given that the technique is largely based on an observer qualitative judgment, experience and examination environment that often vary depending on the facility infrastructure including regional resources leading to high rate of false positivity and low specificity [9].

A more promising alternative for rapid, low-cost, and high-volume HPV testing in low and middle income Countries (LMICs) is the use of Immunostain p16/Ki67 rather than the traditional VIA [10]. The dual stain p16/Ki67 have been shown to be an efficient diagnostic triage test in a number of clinical trials [11,12]. p16INK4a protein is derived from the host p16INK4a /CDKN2A tumor suppressor gene found at chromosome 9 [13]. In humans, this cytoplasmic antigen has been identified as a biomarker for transforming HPV infection and therefore can be used as a surrogate marker of hr-HPV infection [13]. Additionally, Ki-67 (MIB-1) is a nuclear antigen proliferative biomarker which is highly expressed in HPV infected mature squamous cells. These usually manifest in a proliferating cell, hence they are closely linked to tumors of the cervix [14]. Collectively, p16 acts as a cell cycle regulatory protein that induces cell cycle arrest, while ki-67 act a cell proliferation marker. Normally; under physiological conditions, the two biomarkers hardly co-express in the same cervical epithelial cells [6]. Thus; co-expression of these two molecules in any cervical cell therefore suggests a deregulation of the cell cycle mediated by hr-HPV infection which predicts the presence of high-grade cervical intraepithelial lesions (HSIL) [15]. Integration of p16ink4a / ki-67 biomarker as a point of care test to be used specifically to identify at one visit cases of cervical dysplasia with subsequent treatment, is an option currently being explored in a number of developing countries [16]; more so with the evolving landscape of HPV vaccination and changes associated with HPV genotype prevalence [17].Results from a recent Rwanda’s screen, notify, see, and treat CC screening program observed that loss to follow-up owing to VIA examination scheduling was a key challenge derailing optimal uptake in sub Saharan Africa ([18]. This study compared the diagnostic performance of p16/Ki-67 cytology with VIA in the detection of cervical precancers.

## Materials and methods

### Study design

We utilized stored provider-collected samples from 189 hr-HPV positive women with colposcopic biopsy in Kenya who participated in a cross-sectional study assessing the influence of co-infections on CIN [19]. The parent study took place between February 2021 and November 2023 at Jaramogi Oginga Odinga Teaching and Referral Hospital and enrolled 517 women aged 25–65 years with cervical abnormalities referred from peripheral facilities in Kisumu County, Kenya (figure 1); after obtaining research ethics approval (ERC-IB/VOL.1/602). Participant sociodemographic and clinical characteristics were collected (table1)’

**Figure 1.**
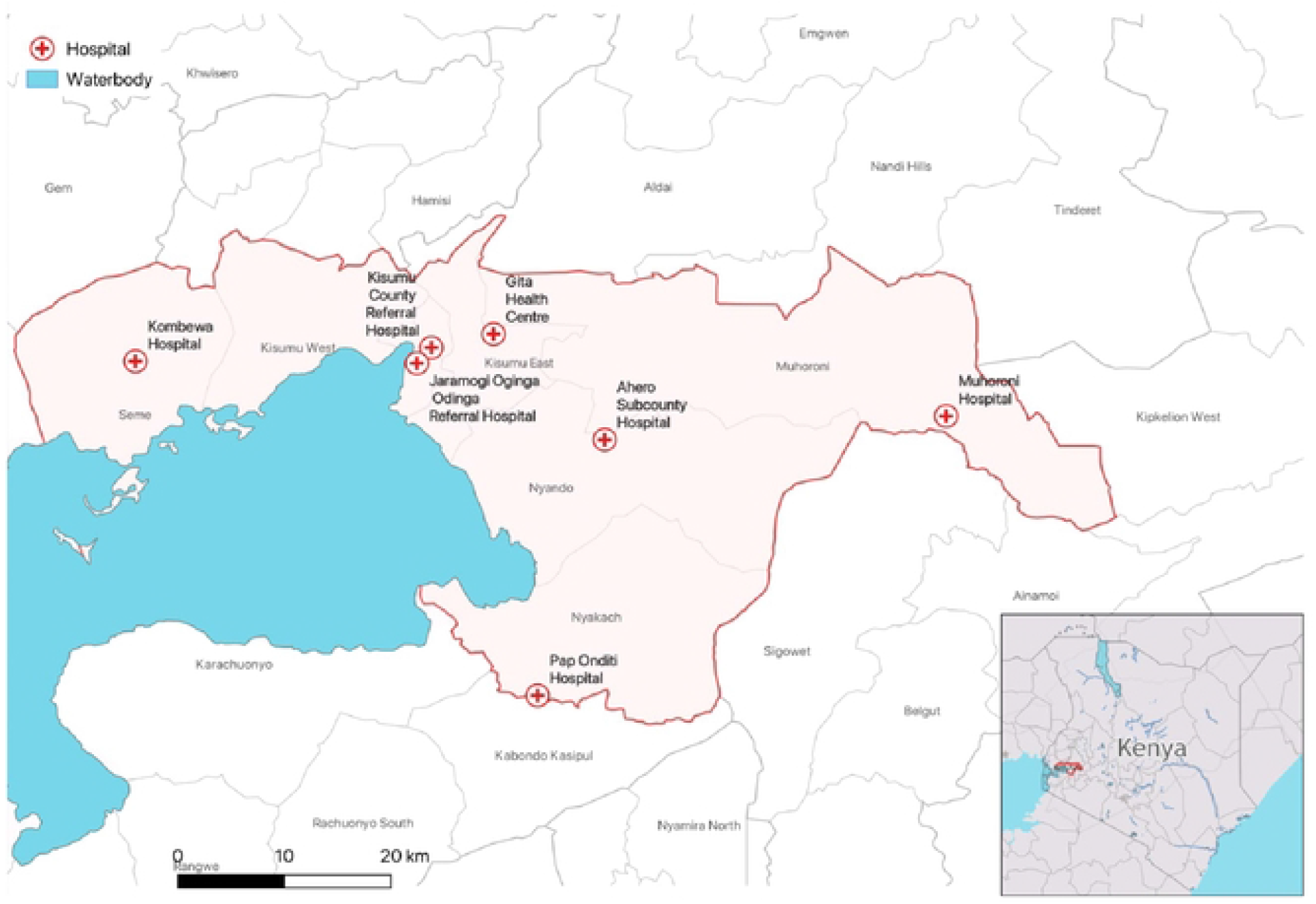
Study map.

**Table 1:** Baseline characteristics of study participants.

| Variables | N=517 | Percentage (%) |
| --- | --- | --- |
| **Age in years** |  |  |
| Age, median (IQR) | 33 | 28,39 |
| Age ≤ 30 | 195 | 37.7 |
| Age > 30 | 322 | 62.3 |
| **Parity** |  |  |
| ≤ 4 children | 315 | 60.9 |
| >4 children | 202 | 39.1 |
| **Marital status** |  |  |
| Married | 456 | 88 |
| Single | 35 | 6.8 |
| Widows | 26 | 5.2 |
| **Contraceptives use** |  |  |
| None users | 50 | 9.7 |
| Implants (Jadele / norplant) | 179 | 35 |
| Injectables (Depo provera) | 202 | 39▪ |
| Hormonal-IUD | 29 | 5.6 |
| oral pills | 57 | 11 |

IUD-intra uterine device; IQR-inter quartile range; majority cases - ▪

Clinical examination involved a gynecological examination with inspection of the cervix uteri and collection of specimens by a gynecologist in a separate room. Cervical sample were collected for HPV DNA and p16/ki-67 cytology test in PreservCyt® Solution (Hologic) using the Cervex-Brush® (Rover). All HPV test positive women, including 80 (42%) VIA positive, 109(58%) VIA negative, 117(62%) p16/ki-67 positive and 72(38%) p16/ki-67 negative women underwent colposcopy and biopsies (if a cervical lesion was present) for disease status verification (table 2). All women were informed about the VIA result immediately, but for laboratory results, they were booked to collect after one month in the next clinic review visit. Cervical biopsies were read by two pathologists at JOOTRH for study quality assurance. Participants who were HPV-positive and VIA positive or p16/ki-67 positive eligible for thermal ablation including those with cervical intraepithelial neoplasia grade 2 or 3 (≥CIN2) on biopsy were treated according to the Kenya national cancer treatment guideline [7]

**Table 2:**
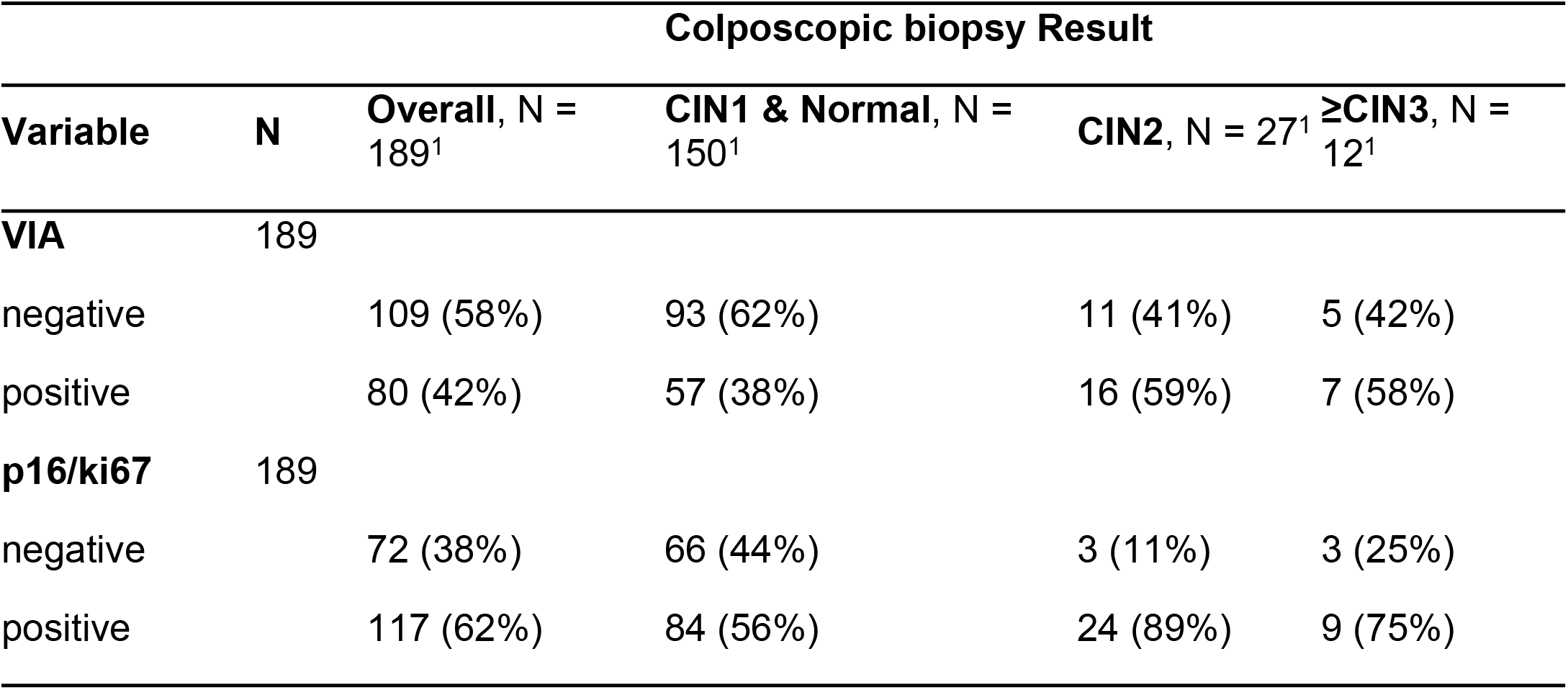
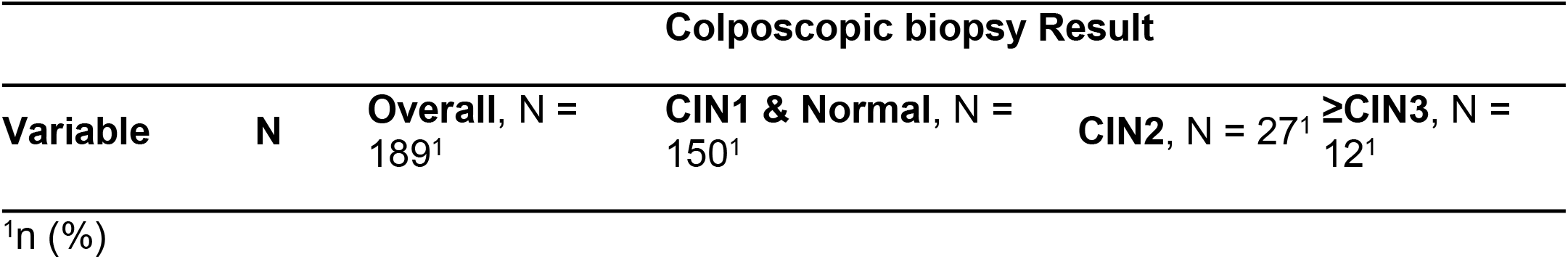
Cervical intraepithelial neoplasia positivity rate by individual test modality.

### HPV-DNA testing

This was performed according to manufacturer’s instruction. HPV DNA was tested using Gene Xpert® HPV (Cepheid, Sunnyvale, California, United States [US]) by transfer of 1-mL aliquot of cervical specimens directly into the Xpert cartridge containing DNA extraction reagents and primers with probes for amplification and HPV detection. Xpert HPV is based on a multiplex real-time PCR targeting E6 and E7 oncogenes of 14 hr-HPV genotypes. The amplification was conducted in five fluorescent channels namely; HPV16, HPV18/45, HPV31/33/35/52/58, HPV51/59, and HPV39/56/66/68, and the results interpreted using Xpert software version 4.8 (Cepheid). HPV positivity was defined if cycle threshold (CT) cutoff was ≤ 40 for HPV16 and HPV 18/45, and ≤ 38 for HPV31/ 35/33/52/58, HPV 51/59, and HPV39/68/56/66. Samples were considered adequate if hydroxymethylbilane synthase was ≤38 cycle threshold.

### p16ink4a / ki-67 Immunostaining test

The remaining residues of PreservCyt specimen after HPV-DNA tests were further subjected to ThinPrep 2000 Processor (Hologic Inc.) to prepare ThinPrep slides which were stained manually using CINtec PLUS p16/Ki-67® by Roche MTM Laboratories according to manufacturer instructions. After primary staining, slides were further incubated with ready-to-use secondary reagents comprising; (a) a polymer reagent conjugated with horseradish peroxidase (HRP) and goat anti-mouse Fab′antibody fragments and (b) a polymer reagent conjugated with alkaline phosphatase and goat anti-rabbit Fab′ antibody fragments. After counterstaining with alcohol-free hematoxylin, a 2-step mounting procedure was applied using an aqueous mounting medium, followed by a permanent mounting medium. The test was performed in the histopathology laboratory of JOOTRH. Interpretation of p16/Ki-67 cytology slides were performed by two cytologists who reviewed all cases for the presence of (double-immune-reactive cells) 1 or more cervical cell (s) showing an evident brown cytoplasmic and a red nuclear staining indicative of p16 and Ki-67 co-expression respectively for a positive result. The same were scanned and shared to a specialist pathologist at Moi teaching and referral hospital for further review. High-grade squamous intraepithelial lesion (HSIL) slides were used as a positive control for each immunoreactions test. The number of p16/Ki-67 Immunostain positive cells and CIN2+ according to VIA examination results were compared. The sensitivity, specificity, positive predictive value (PPV) and negative predictive value (NPV) of the two tests modalities were calculated using SPSS software (version 25.0). Estimates were provided with 95% CIs. The significant differences in sensitivity, specificity, PPV, and NPV were calculated using the exact McNemar χ2 test. Statistical significance was defined as p < 0.05. All statistical test performed were two-sided.

## RESULTS

### Performance of p16/ki-67 cytology and VIA

Table 2 and figure 2 show the overall positivity between p16/ki-67 Immunostain and VIA examination on the 189 stored specimens. Of these 189 specimens, 80 (42%) and 117(82%) were CIN positive on VIA and p16/ki-67, respectively. There were 39 cases of pathologically confirmed ≥CIN2, including 27 cases of CIN2 and 12 cases of ≥CIN3. There were 150 cases defined as CIN1; including 56 cases of CIN1 and 94 cases of normal pathology. The sensitivity, specificity, PPV, NPV, and accuracy of p16/Ki-67 cytology and VIA examination for detection of ≥CIN2 are shown in (table 3). The sensitivity of p16/Ki-67 cytology was 84.6%, which was significantly higher than that of VIA examination (59.0%, p = 0.024). The specificity of p16/Ki-67 cytology and VIA examination were 44.0% and 62.0% (p = 0.003), respectively. The PPV of p16/Ki-67 dual-stain was 28.2%, which was comparable to VIA examination (28.8%, p = 0.025). The NPV of p16/Ki-67 dual-stain was 91.7%, which was significantly higher than that of VIA examination (85.3%, p = 0.002).

**Table 3:** Performance of p16INK4a/Ki-67 and VIA tests to detect ≥CIN2.

| Colposcopic-biopsy | P16/ki-67 test |  | VIA test |  |  |
| --- | --- | --- | --- | --- | --- |
|  | n/No. | %(95%CI) | n/No. | %(95%(CI) | p-value |
| Sensitivity | 33/39 | 84.6 (69.5 – 94.1) | 23/39 | 59.0 (42.1 – 74.4) | 0.024 |
| Specificity | 66/150 | 44.0 (35.9 – 52.3) | 93/150 | 62 (53.7– 69.8) | 0.003 |
| PPV | 33/117 | 28.2 (21.7 – 36.3) | 23/80 | 28.8 (23.1 – 34.5) | 0.025 |
| NPV | 66/72 | 91.7 (87.8 – 94.7) | 93//109 | 85.3 (80.0 – 89.7) | 0.002 |

≥ CIN2, cervical intraepithelial neoplasia grade 2 or worse; 95%CI, 95 percent confidence interval; % proportions. VIA, visual inspection with acetic acid; PPV, positive predictive value; NPV, negative predictive value.

**Figure 2.**
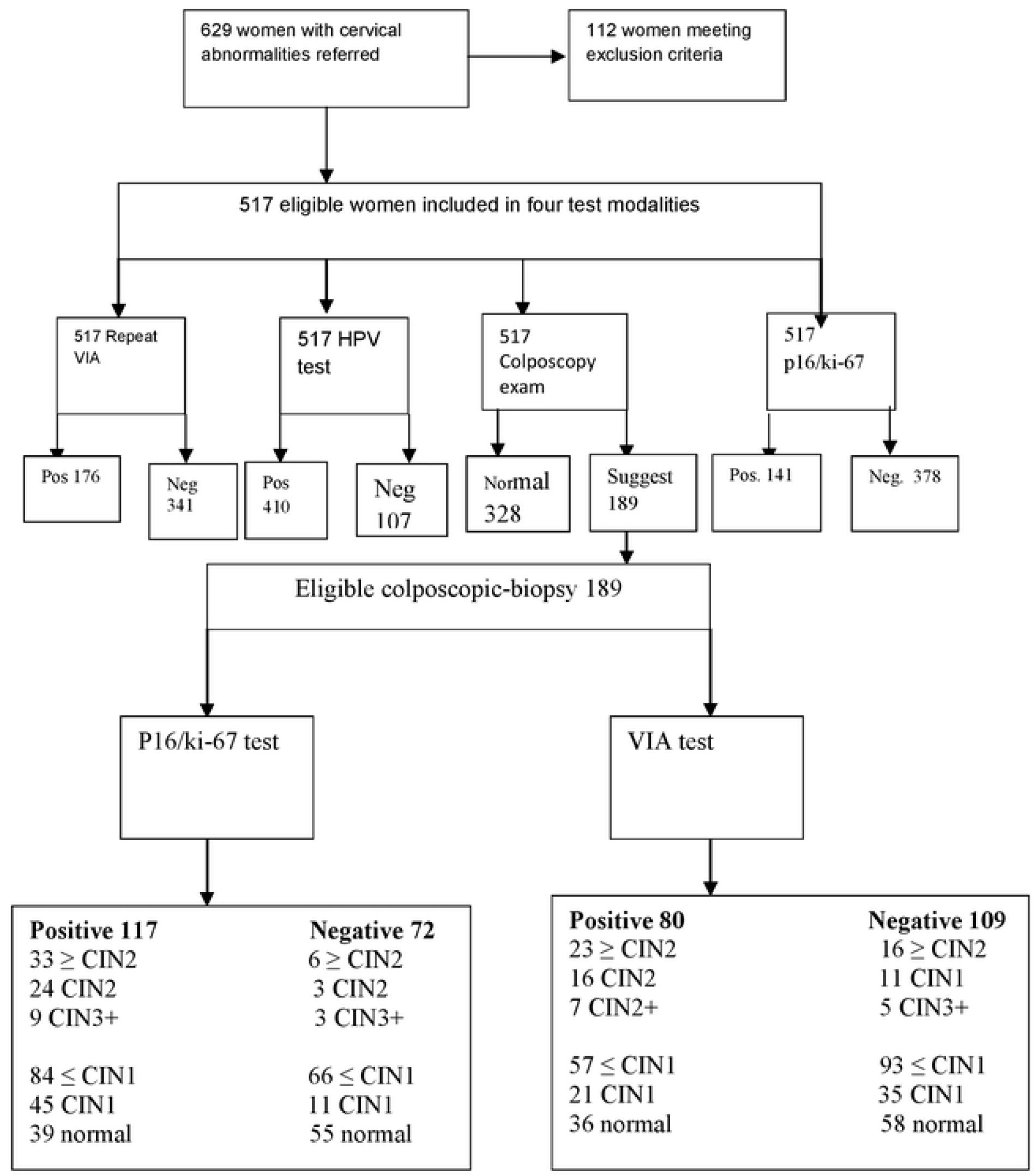
The flowchart.

## Discussion

In an effort to eliminate cervical cancer disease in the globe, the World Health Organization (WHO) recently unveiled new screening and treatment guideline intended to accelerate and promote faster and better interventions outcome as a matter of public health priority in what was dubbed “see and treat” strategy [2]. Among recommendation in the guideline is the implementation of DNA-based HPV testing as a primary screening tool which many countries including Kenya have adopted by gradually transitioning from Pap cytology testing to HPV-based screening or co-testing [7]. So far, major strides have been realized following the adoption of HPV-DNA testing as a primary tool with subsequent triage using VIA where Pap cytology is not available [7]. Notwithstanding the milestones, the low intra-observer agreement associated with VIA has been noted in a number of studies locally [20,21], and in some parts of Africa [8]; thus, prompting a call for a more efficient alternative triage tool. In our study, only 20.6% (39 /189) of all high-risk HPV-positive women who underwent colposcopy and biopsy had ≥CIN2 as shown in figure 2, suggesting that low specificity associated with HPV-DNA tests could lead to a high referral rate for colposcopy and cervical biopsy as reported earlier [14,22]. Accordingly, the need to develop a more efficient triage test for hr-HPV-positive women in order to reduce unnecessary colposcopic referrals can never be overemphasized.

In this study, we compared the performance of p16/Ki-67 cytology and VIA examination in the triage of primary HPV screen-positive women. For the detection of cervical intraepithelial neoplasia 2 and above (≥CIN2), the sensitivity of p16/Ki-67 cytology was significantly higher than VIA examination (84.6% vs. 59.0%, p = 0.024), whereas the specificity remained comparable to VIA examination (44.0% vs. 62.0%, p = 0.003 respectively). Majority of the previous studies conducted on p16/Ki-67 cytology for the prediction of ≥CIN2 yielded similar sensitivity; thus, supporting its potential as a promising and efficient triage tool for high-risk HPV-positive women [11,14,22]. In our systematic review of the same study [23], the reported test performance and the receiving operating characteristics curves conducted from several large scale longitudinal and prospective studies all agreed that implementing p16/ki-67 assay as a triage for HPV positive women to be used at one visit with subsequent treatment is feasible. Similar investigations from China, Romania, Singapore, Slovenia and Denmark analyzing the performance of p16/ki-67 cytology with HPV-DNA test, Pap cytology or Colposcopy recorded higher sensitivity range of 66.0% - 96.7% and specificity range of 51.6% - 93.0% for p16/ki-67 as compared to the rest of the tests showing sensitivity range of 42.1% - 85.7% with specificity range of 14.7% - 95.2% [24,25]. Beyond its potential as an alternative co-test for primary HPV positive women, the assay can also be useful in predicting the regression or progression of CIN2 [16]; and as well serve as a triage tool for younger women aged ≤30 years old presenting with ASC-US or LSIL [26]. Although variable sensitivities have also been reported in some published research [22], this study maintain that the observed variation could possibly be due to some technical problems caused by insufficient cellularity [11], including the variation in study populations. In this study, we included subgroup of women referred for abnormal cytology; thus, explaining the relatively lower specificity observed both in p16/ki-67 cytology and VIA examination. Moreover, both tests showed lower but similar positive predictive value (PPV) in the detection of high grade ≥CIN2, (28.2% vs. 28.8%, p =0.025 respectively), although p16/ki-67 recorded a significantly higher negative predictive value than VIA (91.7% vs 85.3%, p = 0.002) which was comparable to previous studies [11,14,27,28], all showing reduced PPV with increased NPVs.

With the ever increasing CC screening coverage through vast community’s outreaches, the beauty of home-based self-sample collection for the primary HPV-DNA test can never be overemphasized considering the cost associated with provider-collected samples, and the challenge of limited resources experienced by many LMIC [3]. Notwithstanding the advantages, including preference by majority women participants; experts have also argued that cellularity of cervical cells in self-collected samples are limited; and may lead to low sensitivity and reliability particularly with p16/ki-67 assay where sample residues from primary HPV testing is preferred for convenience [11]. Consequently, women testing hr-HPV positive on self-collected samples may still be required to re-visit their clinicians for additional provider-collected cervical smear; thus, pausing a challenge to this immunostaining technology [11] and to the model of “see and treat program” [18], particularly in LMICs where same-day treatment decisions are often necessary after primary screening to reduce patient loss-to-follow-up [29]. This study had some limitations. First, the participants were recruited from a gynecological clinic having been referred from peripheral facilities with either vaginal or cervical abnormalities. This facility-based recruitment, coupled with structural inefficiency in some rural setting that conveyed referrals limit the generalization of the study findings. Secondly, although HPV genotyping is ideal in determining the specific genotypes promoting CIN prevalence in the region, this was not possible owing to the diagnostic platform employed.

## Conclusion

This study evaluated the diagnostic efficacy of p16/ki-67 cytology as a possible alternative triage for HPV positive women. The Immunostain showed higher sensitivity in detecting high grade CIN 2 and above (≥CIN2) with a specificity comparable to VIA. Accordingly, these results suggest that p16/ki-67 cytology is superior to VIA in identifying high-grade CIN; and represents a promising approach as an efficient triage strategy for high-risk HPV-positive women.

## Data Availability

All relevant data are within the manuscript and its Supporting Information files

## Abbreviations

CI: confidence interval
CC: Cervical Cancer
CIN 1, 2, 3: Cervical Intraepithelial Neoplasia (Grade 1, Grade 2, Grade 3)
ICO-IARC: Catalan Institute of Oncology / International Agency for Research on Cancer
HPV: Human Papilloma Virus
hr-HPV: High Risk Human Papilloma Virus
JOOTRH: Jaramogi Oginga Odinga Teaching and Referral Hospital.

## Supplementary Information

The dataset used or analyzed during the current study are available from the corresponding author on reasonable request

## Acknowledgments

We thank our study participants and staffs of JOOTRH, Kenya for their participation in sample collection and providing institutional approval of the study. Much thanks to the Association of Kenya Medical Laboratory Scientific Officers (AKMLSO) for organizing workshops and scientific conferences that brought together both local and international partners some of whom provided technical skills, expertise and technologies useful in this study. We equally appreciate Sustainable Development for Health on HIV (SD4H) program of Maseno University for supporting review of manuscript through organized workshop mentorship program courtesy of Fogarty International Center of the National Institute of Health (NIH) under Award Number D43TW011306; particularly in statistical data analysis as provided by Prof. Lucas Othuon. More thanks to Dr. Thomas Ongalo and Mr. Felix Humwa for additional statistical analysis

## Author contributions

CGO1, LO1, and BG1conceived the research topic and design CGO1, and LO1 participated in acquisition, analysis and interpretation of data, CGO1, LO1, BG1, reviewed and approved the manuscript.

## Availability of data and materials

The datasets used or analyzed during the current study are available from the corresponding author on reasonable request

## Declarations

### Ethics approval and consent to participate

The institutional review board of JOOTRH provided ethics approval of the study number ERC.IB/VOL.1/602. Written informed consent was obtained from all participants.

### Consent for publication

Consent was not needed as we used de-indentified data.

